# Neutralizing antibodies to Omicron after the fourth SARS-CoV-2 mRNA vaccine dose in immunocompromised patients highlight the need of additional boosters

**DOI:** 10.1101/2022.11.19.22282537

**Authors:** Maria Rescigno, Chiara Agrati, Carlo Salvarani, Diana Giannarelli, Massimo Costantini, Alberto Mantovani, Raffaella Massafra, Pier Luigi Zinzani, Aldo Morrone, Stefania Notari, Giulia Matusali, Giuseppe Lauria Pinter, Antonio Uccelli, Gennaro Ciliberto, Fausto Baldanti, Franco Locatelli, Nicola Silvestris, Valentina Sinno, Elena Turola, VAX4FRAIL study Group, Maria Teresa Lupo Stanghellini, Giovanni Apolone

## Abstract

Immunocompromised patients have been shown to have an impaired immune response to COVID-19 vaccines. Here we compared the B-cell, T-cell and neutralizing antibody response to WT and Omicron BA.2 SARS-CoV-2 virus after the fourth dose of mRNA COVID-19 vaccines in patients with hematological malignancies (HM, n=71), solid tumors (ST, n=39) and immune-rheumatological (ID, n=25) diseases. We show that the T-cell response is similarly boosted by the fourth dose across the different subgroups, while the antibody response is improved only in patients not receiving B-cell targeted therapies, independent on the pathology. However, 9% of patients with anti-RBD antibodies did not have neutralizing antibodies to both virus variants, while an additional 5.7% did not have neutralizing antibodies to Omicron BA.2, making these patients particularly vulnerable to SARS-CoV-2 infection. The increment of neutralizing antibodies was very similar towards Omicron BA.2 and WT virus after the third or fourth dose of vaccine, suggesting that there is no preferential skewing towards either virus variant with the booster dose. The only limited step is the amount of antibodies that are elicited after vaccination, thus increasing the probability of developing neutralizing antibodies to both variants of virus. Hence, additional booster doses are recommended to frail patients.

## Introduction

Vaccination against SARS-CoV-2 has saved millions of lives in populations at risk of developing severe COVID-19 disease (i.e. above 60 years, with comorbidities or immunocompromised patients) (Barouch, 2022). It is estimated that in the period from March to December 2021, only in Colombia, vaccination has avoided 32.4% of expected deaths of COVID-19 in individuals older than 60 (Rojas-Botero et al., 2022). A mathematical model has predicted a reduction of 60% of deaths in one year, a percentage which changes according to vaccine coverage, meaning 14.4 millions of deaths avoided globally (Watson et al., 2022). Vaccination schedules generally include two vaccinations plus a booster dose. Indeed, we have recently described that a third dose of SARS-CoV-2 mRNA vaccination is important to augment anti-spike SARS-CoV-2 neutralizing antibodies and T cell responses, but in some immunocompromised individuals it is still not sufficient to reach antibody titers similar to those of healthy individuals (Corradini et al., 2022). A systematic metanalysis has confirmed this data taking into consideration 82 studies of which 77 based on mRNA vaccination schedules (Lee et al., 2022). A fourth dose is thus likely to benefit immunocompromised patients. However, in some conditions such as common variable immunodeficiency (CVID) while the third dose definitely improved their antibody titers, a fourth dose was only slightly improving the response (Nielsen et al., 2022). If the study included a higher number of patients (currently n=33) there could have been a statistically significant improvement of the response, but the possibility that in these patients it is difficult to overcome the immunocompromised condition remains. In another report, although the fourth dose raised the level of neutralizing antibodies from 80 to 96% at one month after the booster in multiple myeloma patients, anti-BCMA (B cell maturation protein) treatment affected the level of neutralizing antibodies after the third and fourth vaccine dose (Ntanasis-Stathopoulos et al., 2022). In this group additional vaccine boosters seemed to even reduce the level of neutralizing antibodies (Ntanasis-Stathopoulos et al., 2022). Similarly, in patients with lymphoid malignancies, anti-B cell therapies have reduced neutralizing antibodies after the fourth dose (Fendler et al., 2022). A second booster dose seems to increase of seven-fold the level of antibodies in 28 patients with systemic lupus erythematosus or rheumatoid arthritis which were not responding to the third dose, without however improving the level of neutralizing antibodies to Omicron variant (Assawasaksakul et al., 2022). Similarly, a second booster dose increased of 9-folds the median antibody level and allowed seroconversion in 60% (3/5) of liver transplanted previously non-responder patients (Harberts et al., 2022) and in 80% of previously low or non-responder hemodialysis patients (Tillmann et al., 2022). The advantage of a fourth vaccine dose in protecting against infection has been recently reported in the healthy population. In Israel, for instance, the fourth dose of vaccine led to a reduction of infections (measured as PCR-positive test result) from 20% in health care workers with three doses to 7% in those with 4 doses, suggesting that the fourth dose correlates with SARS-CoV-2 infection protection, even to Omicron (Cohen et al., 2022). An increase in antibody levels and T cell response after the fourth dose (Bar-Haim et al., 2022), was observed without however major adverse events (Romero-Ibarguengoitia et al., 2022). In long term care facility resident populations, vaccine efficacy increased with every received dose reaching a protection of 49% against infection, 69% against symptomatic infection, and 86% against severe disease after the fourth dose in Canada (Grewal et al., 2022); and in Israel, after the fourth dose, there was a reduction of 34% against overall infection, 64% hospitalizations for mild-to-moderate illness, 67% against severe illness, and 72% against related deaths during the Omicron variant surge (Muhsen et al., 2022). When individuals older than 60 were analyzed the fourth dose reduced of 32% the hospitalization and 22% COVID-19 related deaths (Arbel et al., 2022). A real-world study on the protection offered by 2, 3 or 4 doses of vaccines confirmed that the booster doses protect against COVID-19 associated hospitalization particularly in individuals older than 50 years (Link-Gelles et al., 2022). This is in line with an increase of antibody titers after the second booster dose in older individuals (Eliakim-Raz et al., 2022). However, this population lacked immunocompromised individuals. Thus, it remains important to understand on a large population whether the fourth dose of mRNA vaccine is beneficial to immunocompromised individuals particularly with respect to associated therapies and whether it induces neutralizing antibodies to the Omicron VOC.

Here we report the follow-up results of VAX4FRAIL, a longitudinal study on COVID-19 vaccination in immunocompromised patients, on the effect of a fourth booster dose of mRNA vaccine on the titer of neutralizing antibodies to both WT and Omicron BA.2 and T cell responses.

## Results

### Patient Characteristics

Between October 2021 and June 2022, 228 patients participating to the VAX4FRAIL study (NCT04848493); received the fourth dose of vaccine. 179 patients received the BNT162b2, 46 the mRNA-1273 vaccine, while for 3 the type of vaccine used was not reported. After having excluded patients resulting positive for antibodies directed to the SARS-CoV-2 nucleocapsid (N) antigen and that presumably had a breakthrough infection after the third or fourth dose, we analyzed the immune response before and after the 4^th^ dose of mRNA vaccine in the remaining 135 patients.

The breakdown of patients receiving the fourth dose of mRNA COVID-19 vaccine was: hematological malignancies (HM, n=71 – 52.6%), solid tumors (ST, n=39 – 28.9%) and immunorheumatological (ID, n=25 – 18.5%) diseases (Table 1). The median age was 63 years (interquartile range 56-71) and 74 patients (54.8 %) were women (Table 1).

**Table 1:** Patients’ characteristics (n=135).

|  | HM (n=71) | ST (n=39) | ID (n=25) | Total (n=135) |
| --- | --- | --- | --- | --- |
| **Gender** (number, %) |  |  |  |  |
| Male | 37 (52.1%) | 15 (38.5%) | 9 (36.0%) | 61 (45.2%) |
| Female | 34 (47.9%) | 24 (61.5%) | 16 (64.0%) | 74 (54.8%) |
| **Age (median, IQR)** |  |  |  |  |
|  | 66 (58-72) | 65 (54-71) | 59 (50-63) | 63 (56-71) |
| **Comorbidities** (number, %) |  |  |  |  |
| Yes | 46 (64.8%) | 27 (69.2%) | 14 (56.0%) | 87 (64.4%) |
| Metabolic | 17 (23.9%) | 9 (23.1%) | 8 (32.0%) | 34 (25.2%) |
| Cardiological | 25 (35.2%) | 13 (33.3%) | 4 (16.0%) | 42 (31.1%) |
| Pneumological | 5 (7.0%) | 4 (10.3%) | 6 (24.0%) | 15 (11.1%) |
| Other | 33 (46.5%) | 19 (48.7%) | 8 (32.0%) | 60 (44.4%) |

### The fourth dose of vaccine is safe and well tolerated

We first evaluated whether the fourth dose of vaccine was well tolerated. We found that the fourth dose was in general well tolerated and the safety profile in line with previous doses (Lupo-Stanghellini et al., 2022) (Table 2). The most common moderate adverse events (AE) were pain at the site of injection (55%), fatigue (42.2%), pain in the bones (36.7%) and headache (26.6%). Very few patients had severe AE which were mostly pain in the bones (9.2%), fatigue (7.3%) and pain at the site of injection (6.4%) as shown in Table 2.

**Table 2:** Vaccine-related reactions at one week after the fourth dose (n=109).

|  | Not at all<br>N (%) | Moderate<br>N (%) | Severe<br>N (%) |
| --- | --- | --- | --- |
| Pain at the injection site | 49 (44.9) | 60 (55.1) | 7 (6.4) |
| Fatigue | 63 (57.8) | 46 (42.2) | 8 (7.3) |
| Headache | 80 (73.4) | 29 (26.6) | 5 (4.6) |
| Bone pain | 69 (63.3) | 40 (36.7) | 10 (9.2) |
| Fever | 90 (82.6) | 19 (17.4) | 4 (3.7) |
| Enlarged lymph nodes | 101 (92.7) | 8 (7.3) | 1 (0.9) |
| Skin rash | 106 (97.2) | 3 (2.8) | - |
| Insomnia | 88 (80.8) | 21 (19.2) | 5 (4.6) |
| Diarrhoea | 96 (88.1) | 13 (11.9) | 1 (0.9) |
| Nausea or vomiting | 98 (89.9) | 11 (10.1) | 2 (1.8) |

### The fourth dose of vaccine improves the humoral and cellular response to the Spike protein

All patient groups displayed a statistically significant improvement of the humoral response after the fourth dose of vaccine (Fig. 1A, P<0.0001 for all the groups). However, the antibody response to the spike RBD protein differed in the disease groups with patients with solid tumors responding better (p<0.001 vs ID and p<0.05 vs HM) followed by hematologic malignancies (p<0.005 vs ID and p<0.05 vs ST) and immunorheumatological diseases (p<0.001 vs ST and p<0.005 vs HM) without major discrepancies when analyzing the patients as a group (Fig. 1A) or individually (Suppl. Fig. 1A). In particular, the only patient in the ST group who did not respond to the third dose became positive (defined as anti-RBD ≥ 7.1 BAU/ml) after the fourth dose (100% of non-responders after the third dose), of the HM patients only 2 out of 21 (9%) negative patients and in the ID group out 3 of 12 (25%) not responding to the third dose responded to the fourth dose. Altogether, 17.6% of the patients not responding to the third dose became positive after the fourth dose.

**Figure 1:**
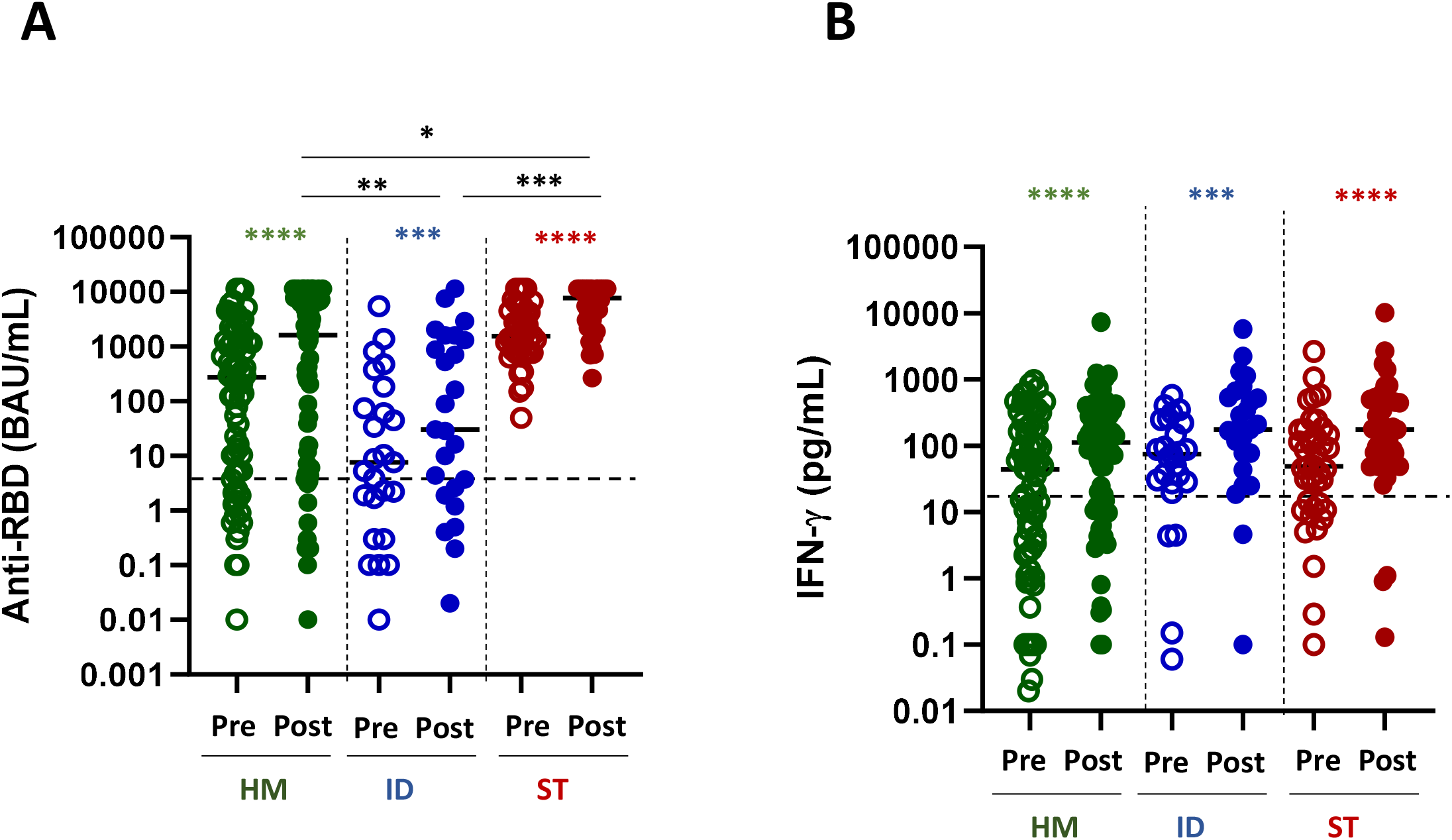
Humoral and T-cell response to the fourth dose of mRNA vaccine in fragile patients. **A**: SARS-CoV-2 specific anti-RBD antibodies (Abs) were measured in sera samples of HM (green dots), ID (blue dots) and ST (red dots) patients before (pre) and after (post) the fourth dose of mRNA vaccine. The level of anti-RBD Abs was expressed as BAU/mL. HM-pre median= 274.8 BAU/mL (IQR 5.2–1357 BAU/mL); HM-post median=1624.0 BAU/mL (IQR 10.7-7269). ID-pre median= 7.6 BAU/mL (IQR 1.0-128.3 BAU/mL); ID-post median= 30.5 (IQR 2.2-1458 BAU/mL). ST-pre median= 1546.0 BAU/ml (810.4-4389 BAU/mL); ST-post median= 7632 (3023-11360 BAU/ml). Differences between anti-RBD Abs titre before and after the fourth dose within the same group were evaluated by Wilcoxon paired test, while differences across groups were evaluated by Kruskal-Wallis. ****P<0.0001; *** P<0.001. **B**: T-cell response were measured in HM (green dots), ID (blue dots) and ST (red dots) patients before (pre) and after (post) the fourth dose of mRNA vaccine. Spike-specific T-cell response was measured after stimulation of whole blood with specific peptides and was expressed as pg/mL of IFN-γ and values 0.12 pg/mL are considered positive. HM-pre median= 44.0 pg/mL (IQR 3.1-185.3 pg/mL); HM-post median=112.6 pg/mL (IQR 14.4-350.9 pg/mL). ID-pre median= 74.8 pg/mL (IQR 28.1-233.0 pg/mL); ID-post median= 175.5 pg/mL (IQR 60.1-595.2 pg/mL). ST-pre median= 48.9 pg/mL (11.9-167.3 pg/mL); ST-post median= 176.0 pg/mL (60.5-495 pg/mL). Differences between anti-RBD Abs titre before and after the fourth dose within the same group were evaluated by Wilcoxon paired test, while differences across groups were evaluated by Kruskal-Wallis. ****P<0.0001; *** P<0.001. HM, hematological malignancies; ID, immune-rheumatological diseases; ST, solid tumors.

Similarly, the spike-specific T-cell response (defined as IFN-γ levels ≥12 pg/mL) was statistically significantly improved in all of the disease groups after the fourth dose (Fig. 1B P<0.0001 for all the groups). Notably, differently from the B-cell response, the T-cell response was very similar across disease groups (no statistically significant differences among groups, Fig. 1B), suggesting that the T-cell response is less dependent on disease or treatment characteristics.

### The presence of neutralizing antibodies to the WT and omicron BA.2 VOC depends on the amount of anti-RBD antibodies elicited after the booster dose

Patients with fragility have been the first to receive a fourth dose of vaccine. The new version of the vaccine targeting omicron VOC was not available at the time of vaccination. Thus, an important question that arises is whether these patients should undergo a booster dose with the adapted versions of the vaccines to the omicron VOC. We therefore evaluated the neutralizing activity of anti-RBD positive sera against SARS-CoV-2 omicron (BA.2) infectivity in a BSL-3 facility. Sera from all anti-RBD positive patients demonstrated a significantly increased neutralizing activity after the fourth dose of vaccine towards the omicron VOC BA.2 (Fig. 2A, P<0.0001 for HM and ST, P= 0.008 for ID). Of note, the neutralization titer after the fourth dose reached higher level in HM and ST than in ID patients (Fig. 2A, HM vs ID, P<0.05; ST vs ID, P<0.001). Similarly to the antibody response, the percentage of anti-RBD positive patients showing a detectable neutralizing activity after the fourth dose of vaccine was higher within ST (97.4%) and HM (80.7%) and lower within ID (66.6%) patients.

**Figure 2:**
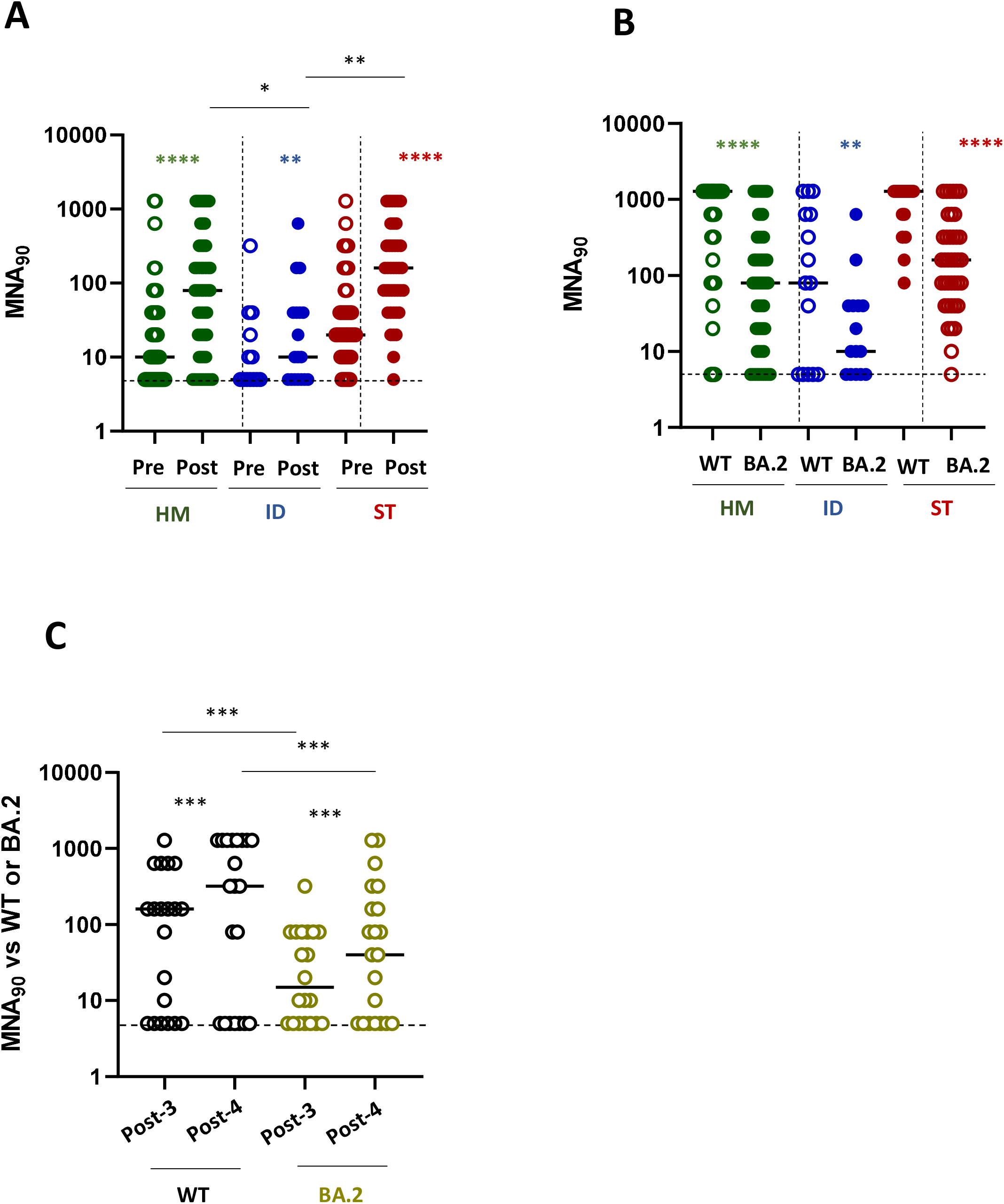
Neutralization response to the fourth dose of mRNA vaccine in fragile patients. **A**: The levels of neutralizing antibodies were quantified by microneutralization assay (MNA_90_) in HM (green dots), ID (blue dots) and ST (red dots) before (pre) and after (post) the fourth dose of mRNA vaccine. The MNA_90_ was measured only in patients showing a positive anti-RBD response. The results were expressed as reciprocal of dilution and values >5 are considered positive. HM-pre median= 10 reciprocal of dilution (IQR 5-40); HM-post median=80 reciprocal of dilution (IQR 10-320). ID-pre median= 5 reciprocal of dilution (IQR 5-30); ID-post median= 15 reciprocal of dilution (IQR 5-40). ST-pre median= 20 reciprocal of dilution (IQR 10-80); ST-post median= 160 reciprocal of dilution (IQR 50-320). Differences between MNA_90_ titre before and after the fourth dose within the same group were evaluated by Wilcoxon paired test, while differences across groups were evaluated by Kruskal-Wallis. ****P<0.0001; ** P<0.01. **B**: The levels of neutralizing antibodies against wild-type (WT) and BA-2 Omicron (BA.2) strains after the fourth dose of mRNA vaccine were compared in HM (green dots), ID (blue dots) and ST (red dots). The results were expressed as reciprocal of dilution and values >5 are considered positive. HM-WT median=1280 reciprocal of dilution (IQR 80-1280); HM-BA.2 median=80 reciprocal of dilution (IQR 10-320). ID-WT median= 80 reciprocal of dilution (IQR 5-640); ID-BA.2 median= 10 reciprocal of dilution (IQR 5-40). ST-WT median= 1280 reciprocal of dilution (IQR 640-1280); ST-BA.2 median= 160 reciprocal of dilution (IQR 40-320). Differences were evaluated by Kruskal-Wallis test. ****P<0.0001; ** P<0.01. **C**: The levels of neutralizing antibodies against wild-type (WT) and BA-2 Omicron (BA.2) strains after the third (post-3) and fourth (post-4) dose of mRNA vaccine were compared in a subgroup of patients (n=21). The results were expressed as reciprocal of dilution and values >5 are considered positive. WT-post-3 median=160 reciprocal of dilution (IQR 5-520); WT-post-4 median= 320 reciprocal of dilution (IQR: 5-1280). BA.2-post-3 median= 15 reciprocal of dilution (IQR 5-80); BA.2-post-4 median= 40 (IQR 5-240). Differences between neutralization titre against WT and BA.2 after the third and fourth dose evaluated by Wilcoxon paired test ***P<0.001. HM, hematological malignancies; ID, immune-rheumatological diseases; ST, solid tumors.

Interestingly, regarding ID patients, only a minority of them displayed a level of neutralization above baseline for the omicron BA.2 VOC. In order to evaluate whether the low level of neutralization titer was evident only towards the omicron VOC or also against the WT SARS-CoV-2, for a reduced number of patients (67 HM, 24 ID and 39 ST), we compared the difference in neutralizing abilities towards the WT and Omicron variant before and after the fourth dose. As shown in Fig. 2B mirroring the increased titer of antibodies induced after vaccination in ST patients, there was a very high level of neutralizing antibodies towards both Omicron BA.2 and WT, with a statistically significantly higher level towards the WT virus (p<0.001). By contrast in HM and even more so in ID patients, the level of neutralization towards both the Omicron BA.2 and WT virus was very low (Fig. 2B). 9% of patients with anti-RBD antibodies did not have neutralizing antibodies to both forms of the virus, while an additional 5.7% did not have neutralizing antibodies only to Omicron BA.2 variant (Suppl. Fig. 2A), making these patients particularly vulnerable to SARS-CoV-2 infection. We supposed that the probability of finding neutralizing antibodies to both forms of the virus depended on the amount of anti-RBD antibodies elicited during the vaccination. Thus we analyzed the correlation between the level of anti-RBD antibody response and neutralizing antibodies to WT or Omicron BA.2. As shown in Suppl. Fig. 2B there was a positive correlation between anti-RBD antibodies and neutralizing antibodies to WT (p<0.0001, R^2^=0.85) or Omicron (p<0.0001, R^2^=0.69). In particular, anti-RBD levels below 100 BAU/mL correlated with no neutralizing antibodies to WT, while the level of anti-RBD correlating with the presence of neutralizing antibodies towards Omicron BA.2 was more difficult to identify. Nevertheless, a level of antibodies above 350 BAU/mL increased the chances of finding neutralizing anti Omicron BA.2 antibodies (86/89, 96.6%), while at levels above 1000 BAU/mL we found only one patient with still no neutralizing antibodies to Omicron BA.2 (Suppl. Fig. 2B). There was also a clear correlation between neutralizing antibodies to WT and Omicron BA.2, suggesting that they may broadly recognize common moieties of the viruses (Suppl. Fig. 2B). Thus, even though patients may have detectable anti-RBD antibodies the probability of finding neutralizing antibodies to Omicron BA.2 is low and correlates on the amount of anti-RBD antibodies.

We then asked if the fourth dose may equally boost the response to the WT and Omicron BA.2 VOC and evaluated the increment of neutralizing antibodies after the 3^rd^ and 4^th^ dose of vaccine towards the two viruses. As shown in Fig. 2C, we found that the fourth dose was inducing a statistically significant (p<0.005) similar increment of neutralizing antibodies towards both viruses (fold increase: 3.29 ± 3.776 for WT and 3.053 ± 3.61 for BA.2). Thus, patients with ID and HM have a reduced neutralizing antibody response to SARS-CoV-2 independent on the variant. Whenever present, this neutralizing response is similarly boosted towards both forms of the virus across different booster doses.

### Anti-CD20 treatment undermines the Response to Vaccination

HM and ID patients often undergo anti-B cell treatment (Rituximab) which can have a major impact on the humoral B cell response to COVID-19 vaccines, as we have shown in this (Corradini et al., 2022) and other cohorts of patients (Azzolini et al., 2022). Thus, humoral and T-cell responses were evaluated in relation to the received or ongoing treatment and its presumed immunosuppression (Table 2). It is clear that B-cell directed treatment has an impact on the anti-RBD response (Suppl. Fig. 3A) but not on the T-cell response (Suppl. Fig. 3B). More in details, in Fig. 3, we analyzed the B- and T-cell response to the fourth dose in relation to underlying disease and treatment. It is clear that both in HM (Fig. 3A, left panel) and ID (Fig. 3B, left panel) the only treatment which strongly affected the B-cell humoral response was anti-CD20 treatment (Rituximab). Patients with HM undergoing anti-CD20 treatment had a reduced anti-RBD response as compared with HSCT (p<0.001), CT (p<0.05) and JAK inhibitors (p<0.01) (Fig. 3A). Patients with ID had a reduced anti-RBD response as compared with the other treatments (p<0.001) (Fig. 3B). By contrast, the different treatments used in solid tumors (ST) had no effect on the induction of the humoral response. As expected, the T cell response to the Spike protein evaluated as IFN-γ production was not affected by anti-CD20 or any other treatment, independent on the pathology of the patients (Figure 3 right panels) Hence, the antibody immune response to the vaccine is dependent mostly on the patients treatment rather than their pathology.

**Figure 3:**
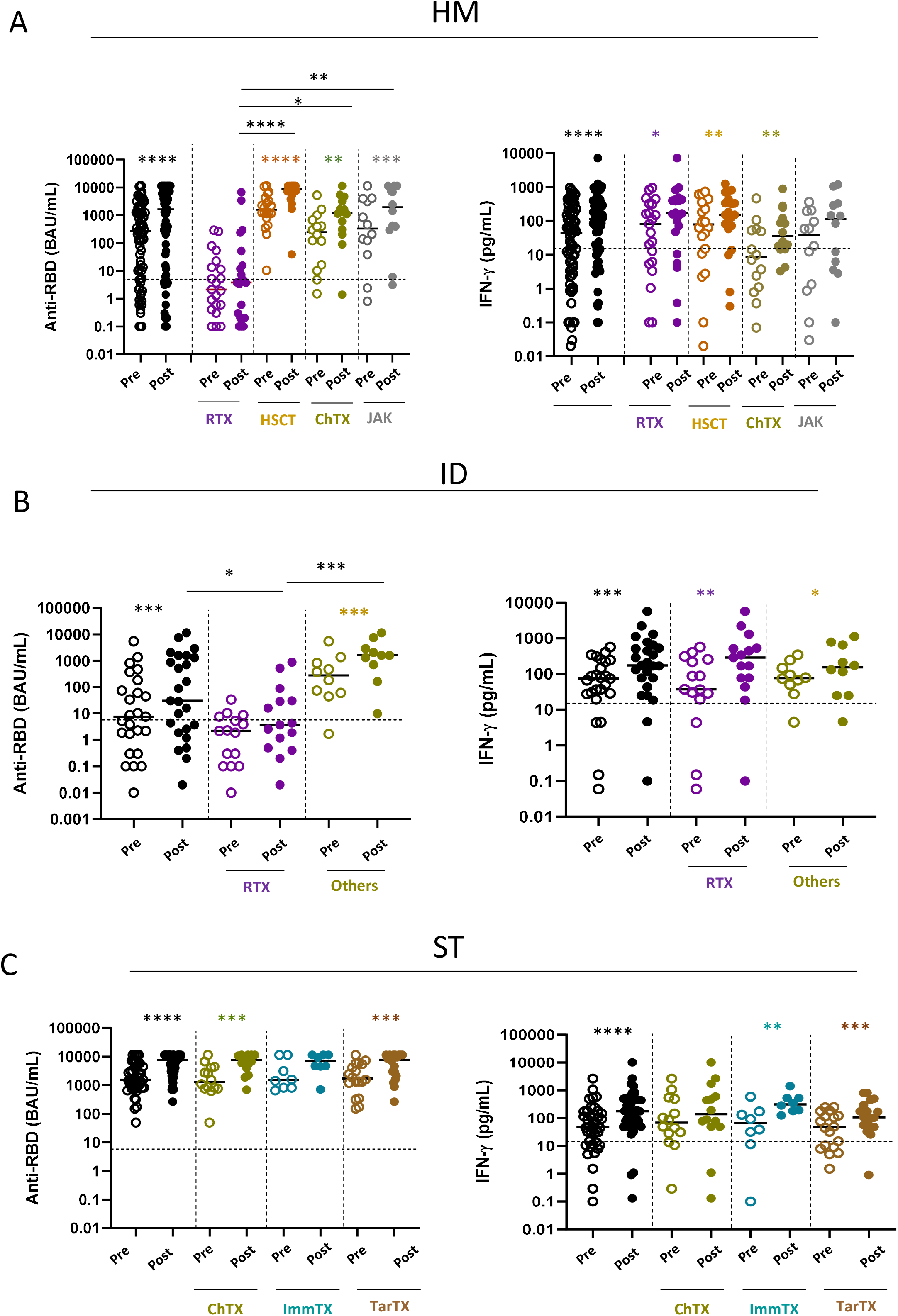
Impact of different treatments on humoral and T-cell response. **A**: SARS-CoV-2 specific anti-RBD Abs and T cell response before (pre) and after (post) the fourth dose of mRNA vaccine were compared in HM patients receiving different treatments: Rituximab (RTX), hematopoietic stem cell transplantation (HSCT), Chemotherapy (ChTX), JAK inhibitors (JAK) or others. **B**: SARS-CoV-2 specific anti-RBD Abs and T cell response before (pre) and after (post) the fourth dose were compared in ID patients receiving different treatments: Rituximab (RTX) and other (methotrexate, mycophenolate mofetil, and azathioprine). **C**: SARS-CoV-2 specific anti-RBD Abs and T cell response before (pre) and after (post) the fourth dose of mRNA vaccine were compared in ST patients receiving different treatments: Chemotherapy (ChTX), Immunotherapy (ImmTX) and Target therapy (TarTX). The median and IQR are described in Supplementary material. Differences between anti-RBD or T cells response level before and after vaccination within the same group was evaluated by Wilcoxon paired test, while differences across groups were evaluated by Kruskal-Wallis. ****P<0.0001, *** P<0.001, **P<0.01, *P<0.05. HM, hematological malignancies; ID, immune-rheumatological diseases; ST, solid tumors.

## Discussion

In this study we show that the fourth dose of SARS-CoV-2 mRNA vaccine is well tolerated and increases the immune response to both WT and Omicron BA.2 VOC. As already reported for the third dose by us and others (Azzolini et al., 2022; Corradini et al., 2022; Lee et al., 2022), the humoral response to the fourth dose was compromised in patients with HM and ID that underwent rituximab (anti-CD20) treatment. However, the fourth dose, induced a similar level of T cell response to the Spike protein in all of the analyzed groups, confirming that fragile patients are capable of mounting a cellular immune response to the vaccine. Interestingly, even if directed to the WT strain of the virus, the vaccine was capable of increasing the level of neutralizing antibodies also to the Omicron BA.2 VOC. The level of neutralization was, as expected, lower than that for the WT version of the virus, however, the increment of neutralizing antibodies after the third and fourth doses was very similar towards both the WT and omicron VOC, suggesting that there is no preferential skewing of the humoral response towards the WT in the booster dose. Interestingly, we observed many patients (9%), particularly those with rheumatologic disorders, whose antibodies were not neutralizing towards both WT and Omicron BA.2, and an additional 5.7% without neutralizing antibodies to Omicron BA.2, and this correlated with the amount of anti-RBD antibodies elicited after the booster dose. This may explain the results of another report on patients with systemic lupus erythematosus or rheumatoid arthritis, where it was observed no neutralizing antibodies to Omicron in patients with an already low response to the third dose (Assawasaksakul et al., 2022). However, it is still remarkable that, even in immunocompromised individuals, the mRNA vaccines elicit antibodies that can recognize very different variants of the virus. This is a characteristic that is less evident during the natural infection (Roltgen et al., 2022). Indeed, the germinal centers reaction after viral infection is limited and thus the immune response is affected in breadth and this may explain why patients that have been infected are easily re-infected by a different VOC and that the immune response to different variants may be compromised (Reynolds et al., 2022; Richardson et al., 2022; Rossler et al., 2022). By contrast, the mRNA vaccines induce a strong germinal center reaction also due to the persistence of the spike protein which is retained in the lymph nodes for up to 2 months after vaccination thus continuously boosting the immune response (Roltgen et al., 2022). We confirm that also in fragile individuals the breadth of the immune response induced by the vaccine is large and includes neutralizing antibodies to the Omicron. We recently demonstrated that anti-Spike IgG can permeate the saliva and thus may protect mucosal sites in the first 3 months after vaccination (Darwich et al., 2022); this correlated with the amount of antibodies present in the blood. Given the low level of neutralizing antibodies against Omicron present in blood, it is likely that an even lower amount is present in the saliva and this may explain why the Omicron was so infectious also in vaccinated people. Consistently, three doses of vaccine are more protective against Omicron than two doses (Chaguza et al., 2022). Patients with cancer are also at higher risk of Omicron than Delta infection, especially patients with HM (Mair et al., 2022) and this may be explained by the low level of neutralizing antibodies to the Omicron VOC. In addition, cancer patients who underwent breakthrough infection with Omicron and had no detectable antibodies to Omicron before, developed these antibodies after infection (Fendler et al., 2022). This may indicate either that even if not detectable, some antibodies to the Omicron were present and probably the B-cells producing them expanded after infection, or, non-mutually exclusive, that T-cell response which is induced also in patients without a detectable B-cell response, may promote a fast induction of the antibody response to Omicron. Either way, these results are encouraging and indicate that vaccination booster doses may indirectly protect against variants of the virus that are not included in the vaccine, even in immunocompromised individuals. Further, it is important to note that anti-CD20 treatment,, as well as other B-cell depleting therapies such as CAR T cells, has the most detrimental effect on vaccination while other treatments used in patients with solid tumors or hemodialysis (Becker et al., 2022) do not affect the fourth dose of vaccination. Finally, the evidence that, regardless of frailties and therapies, the T-cell compartment can respond effectively to vaccination represents an important observation, as antigen specific T cells are actively involved in the protection against the severe disease (Brasu et al., 2022). This also prompts patients undergoing anti-CD20 therapies to proceed to vaccination even if their B-cell response will not be detectable.

In conclusion, we show that the fourth dose of vaccine increases the level of circulating anti-RBD antibodies in all of the patients. These levels however remain low in patients undergoing B cell targeted therapies. Nevertheless, the T-cell response is boosted also in immunocompromised patients undergoing anti-CD20 treatment, thus providing a reinforced line of protection against the severe disease. Similarly to healthy individuals, in fragile patients the breadth of antibodies is ample and includes antibodies recognizing WT and to a much lower extent the Omicron BA.2 VOC. Thus, it is advisable for immunocompromised patients to undergo a 5^th^ dose of vaccine to further boost the development of an Omicron-directed B-cell response or to boost the T-cell response in anti-CD20 treated patients.

### Limitations of the study

During the course of the study, the policy of vaccination has changed in Italy and only some of the centers of the VAX4FRAIL study protocol could administer the fourth dose. Thus, we could not follow-up with the population of patients with neurological disorders and some of the patients with the other pathologies. Still we had enough patients to draw conclusions from the fourth dose. We have tested the neutralization to BA.2 and not BA.4 or BA.5 Omicron VOC. Hence, the conclusions are not generalizable to the most recent VOC which seem to escape the immune response elicited by the WT vaccine (Bowen et al., 2022; Cao et al., 2022)

## Methods

### Study Design

The study was approved by the Italian Medicines Agency (AIFA) and by ethics committee (code 304, 2021) and written informed consent was obtained from all the participants. The inclusion criteria were age ≥18 years, SARS-CoV2 mRNA-based vaccination (fourth dose) and a life expectancy of at least 12 months at the time of vaccine administration. The main exclusion criterion was the presence of a previous laboratory-confirmed SARS-CoV-2 infection (documented by serology and/or molecular test). Moreover, patients experiencing a molecularly confirmed SARS-CoV-2 breakthrough infection (RT-PCR assay) or seroconversion to anti-Nucleocapsid antibody during follow-up were also excluded (n=93).

### Laboratory Procedures

Anti-spike SARS-CoV-2 humoral (binding and neutralizing antibodies) and T-cell response were monitored before 4^th^ dose vaccination (pre-4) and after 10-20 days after vaccination (post-4) in each group. Moreover, the immune response was also compared in treatment-specific subgroups.

The response to vaccination was assessed by quantifying the anti-Nucleoprotein-IgG (anti-N) and the anti-RBD-IgG (Architect® i2000sr Abbott Diagnostics, Chicago, IL). Anti-N IgG were expressed as a ratio (S/CO) and values were considered positive when ≥ 1.4. Anti-RBD-IgG were expressed as binding arbitrary units (BAU)/mL and values were considered positive when ≥ 7.1. The anti-N quantification was aimed to identify asymptomatic SARS-CoV-2 infections during the follow-up and to exclude these patients from the analysis. The median level of anti-RBD was calculated using values from all patients (both non-responder and responder patients). A neutralization assay against BA.2 variant was performed on anti-RBD positive samples to evaluate the functional activity of vaccination-induced antibodies against Omicron. In a subgroup of patients (n=71), the neutralization titer against the Wild Type SARS-CoV-2 strain was also evaluated (22 HM, 10 ID and 39 ST).

The cell-mediated immune response to vaccination was assessed through a standardized whole blood assay (Agrati et al., 2021), shared between clinical centers. Peripheral blood was collected in heparin tubes and stimulated with a pool of peptides spanning the Spike protein (Miltenyi Biotech, Germany) at 37°C (5% CO2). A pool of S protein peptides was generated by combining the PepTivator® SARS-CoV-2 Prot_S, the PepTivator® SARS-CoV-2 Prot_S1 and the PepTivator® SARS-CoV-2 Prot_S+ (all by Miltenyi Biotech) and was used for cell stimulation at the final concentration of 0.1 ug/ml. The spontaneous cytokines release was calculated in unstimulated culture (background) and a superantigen (SEB) was used as positive control. Plasma was harvested after 16-20 h of stimulation and stored at −80°C. IFN-γ (detection limit 0.17 pg/ml) was quantified in the plasma samples using an automatic ELISA (ELLA, Protein Simple). The results are expressed as the amount of each cytokines after subtracting the background. A concentration equal to or greater than 12 pg/mL of IFN-γ was considered as positive.

### Statistical Methods

Quantitative variables were summarized as median and interquartile range (IQR), while categorical variables were reported as absolute count and percentage. Differences in seroconversion rates across subgroups were analyzed using the chi-square test, and from a multivariable logistic regression model we obtained the odds ratios with their 95% confidence intervals (CI). The model outcome was the seroconversion status after the fourth dose (yes vs no) and independent variables were identified on the basis of availability as required by the study protocol (disease, age, gender, comorbidities and therapy) and used to adjust the vaccination effect on outcome. Current therapy was stratified as follows: for HM patients: Rituximab (RTX), hematopoietic stem cell transplantation (HSCT), chemotherapy (CT), JAK inhibitors (JAK); ID patients: anti-CD20 (Rituximab, RTX), and others (methotrexate, mycophenolate mofetil, and azathioprine); ST patients: chemotherapy (CT), Immunotherapy (ImmTx) and targeted therapy (TarTx).

The Wilcoxon test was used to assess differences between pre-4 and post-4, while Mann-Whitney test was used to assess differences in antibody titers across groups.

The SPSS v.20.0 (IBM) statistical software was used for the analysis.

## Data Availability

All data produced in the present study are available upon reasonable request to the authors

## Supplementary files

**Supplementary Figure 1: SARS-CoV-2 specific anti-RBD Abs per individual patient**.

Anti-RBD were measured in sera samples of HM (green dots), ID (blue dots) and ST (red dots) patients before (pre) and after (post) the fourth dose of mRNA vaccine. The level of anti-RBD Abs was expressed as BAU/mL. Differences between anti-RBD titre before and after vaccination were evaluated by Wilcoxon paired test. ****P<0.0001, *** P<0.001.

HM, hematological malignancies; ID, immune-rheumatological diseases; ST, solid tumors.

**Supplementary Figure 2: Cross reactivity of anti-RBD Abs induced by vaccination**.

**A**: The number (percentage) of patients showing a negative or positive anti-RBD response (cut-off 7.2 BAU/mL) after the fourth dose of mRNA vaccine is shown. Patients with a positive anti-RBD response were further divided on the basis of their neutralization capability against WT and BA.2 viral strains.

**B**: The correlation between the levels of anti-RBD Abs and neutralization titre (WT or BA.2) after the fourth dose as well as the correlation between the neutralization titre against WT and BA.2 viral strains are shown. Each black dot represents one sample. The analysis was performed by using the Spearman test and Rho and p values are indicated in the figure.

**Supplementary Figure 3: Impact of Rituximab therapy on humoral and T cell response**

**A**: SARS-CoV-2 specific anti-RBD Abs before (pre) and after (post) the fourth dose of vaccine were compared in all enrolled patients receiving Rituximab (RTX) or other treatments.

All-pre median= 462.5 BAU/mL (IQR: 10.4-1913 BAU/mL); all-post median= 2212 BAU/mL (IQR: 51.6-8391 BAU/mL). RTX-pre median= 2.1 BAU/mL (IQR: 0.3-10.0 BAU/mL); RTX-post median= 3.7 BAU/mL (IQR: 0.3-30.1). Other-pre median=1155 BAU/mL (IQR: 316.4-3145 BAU/mL); Other-post median=5446 BAU/mL (IQR: 1537-11360 BAU/mL)

**B**: SARS-CoV-2 specific T cell response before (pre) and after (post) the fourth dose of vaccine were compared in all enrolled patients receiving Rituximab (RTX) or other treatments.

All-pre median= 49.5.0 pg/mL (IQR: 8.96-177.1 pg/mL); all-post median= 147.0 pg/mL (IQR: 46.7-439.1 pg/mL). RTX-pre median= 63.0 pg/mL (IQR: 7.9-298.7 pg/mL); RTX-post median= 171 pg/mL (IQR: 53.9-475.0 pg/mL). Other-pre median= 49.0 pg/mL (IQR: 8.9-148 pg/mL); Other-post median= 124.5 pg/mL (IQR: 32.6-345.0 pg/mL).

