## Supplementary figures and images for "Neutralizing antibodies to Omicron after the fourth SARS-CoV-2 mRNA vaccine dose in immunocompromised patients highlight the need of additional boosters"

### Supplementary Figures

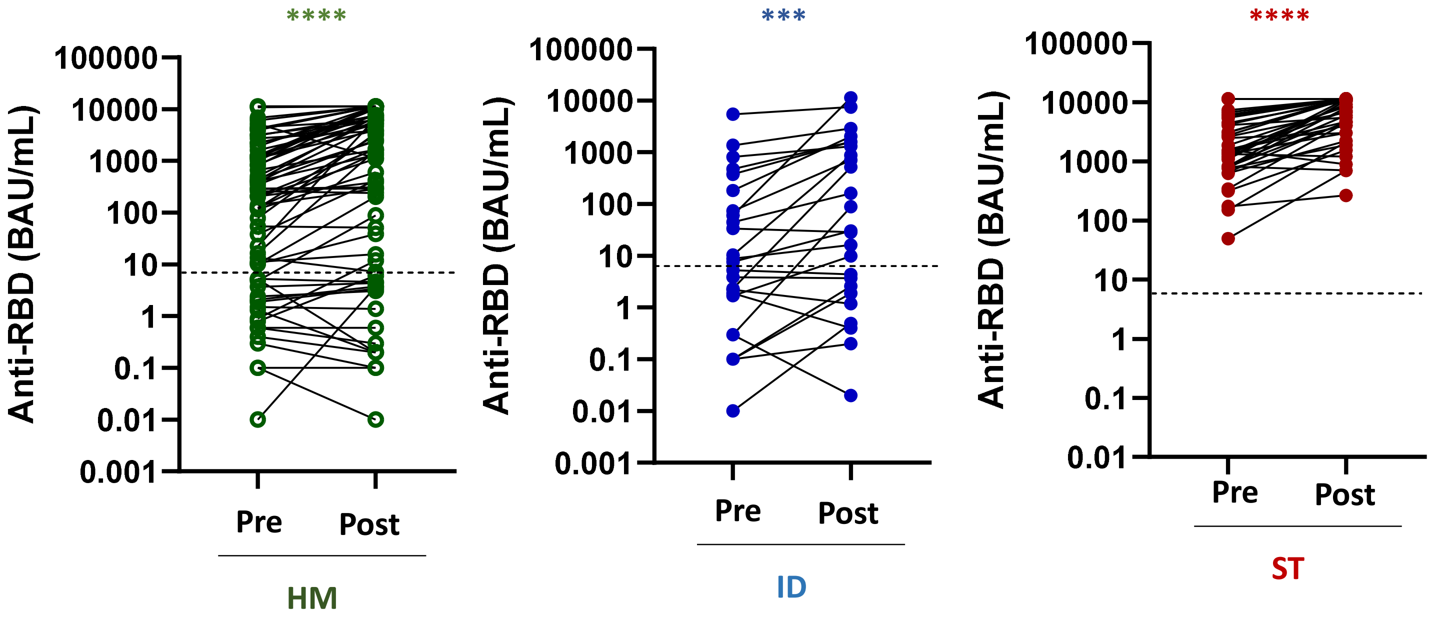

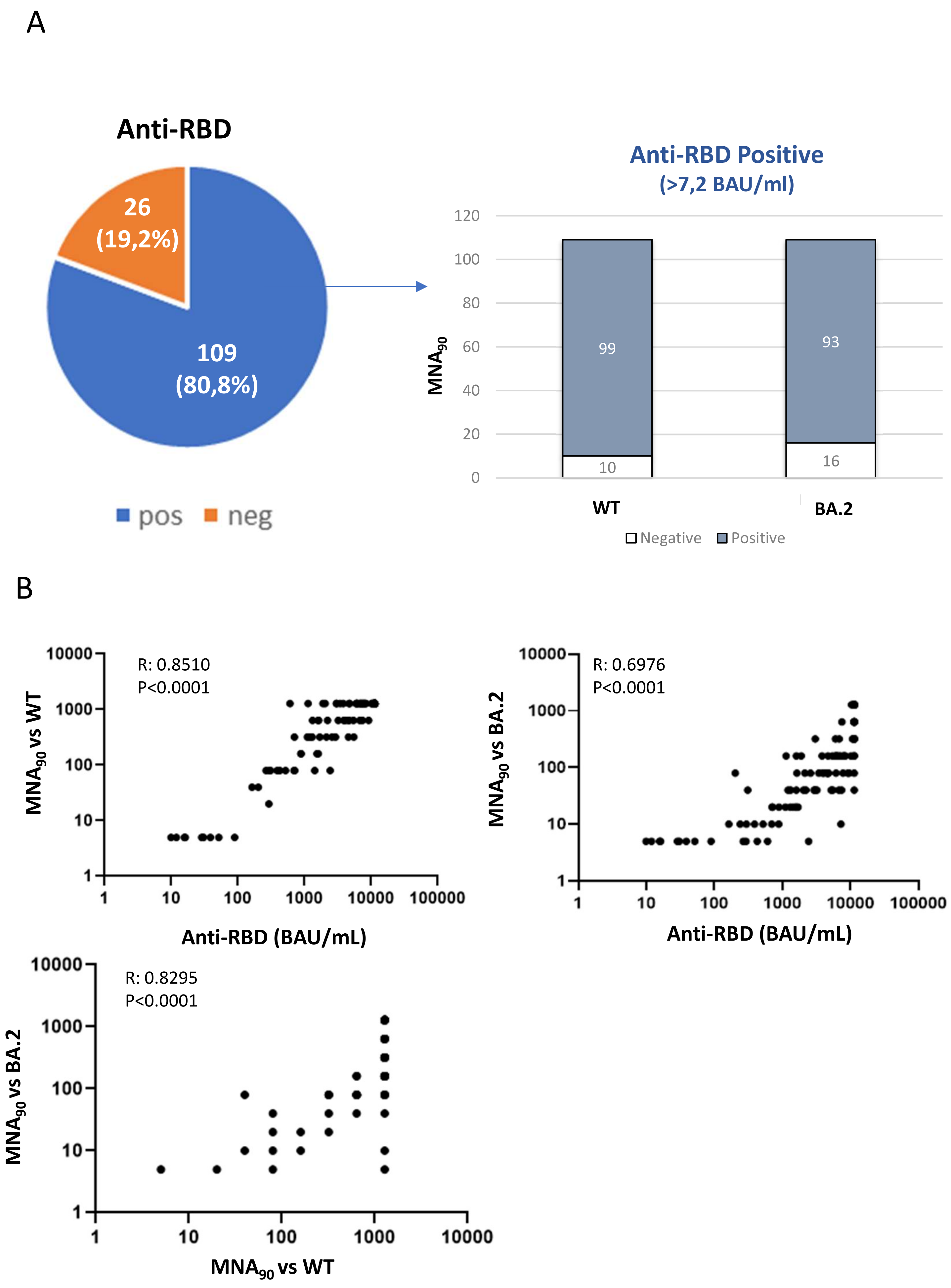


Supplementary Figure 2

A

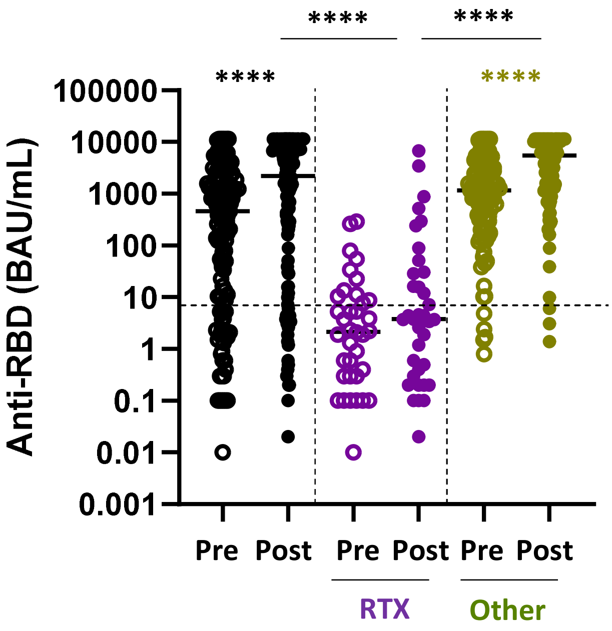


B

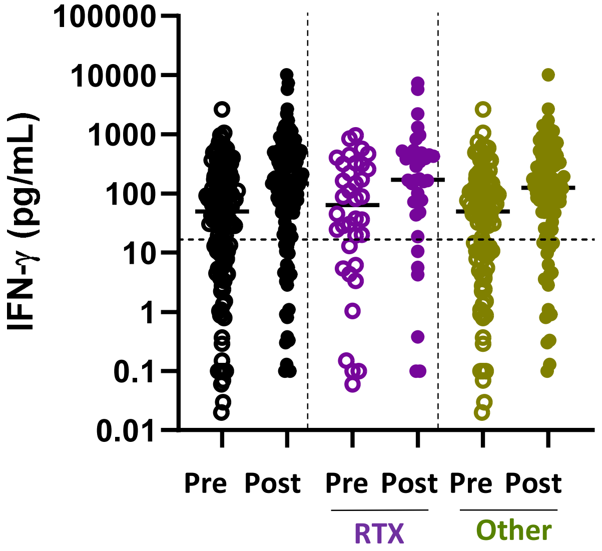


Supplementary figure 3
