## Supplementary Materials for "Neutralizing antibodies to Omicron after the fourth SARS-CoV-2 mRNA vaccine dose in immunocompromised patients highlight the need of additional boosters"

**Impact of different treatments on humoral and T-cell response** (Figure 3)

The medians and interquartile ranges (IQR) of humoral (Anti-RBD) and T cells response in patients with hematological malignancies (HM), immune-rheumatological diseases (ID) and solid tumors (ST) before and after the fourth dose of mRNA vaccine are detailed in the Tables below. Enrolled patients were divided on the basis of the treatment. RTX: Rituximab; HSCT: Hematopoietic stem cell transplantation; ChTX: Chemotherapy; JAK: Jak inhibitors; ImmTX: Immunotherapy; TarTX: Target Therapy.

**A: HM patients**

|  | **All**  **median (IQR)** | | **RTX**  **median (IQR)** | | **HSCT**  **median (IQR)** | | **ChTX**  **median (IQR)** | | **JAK**  **median (IQR)** | |
| --- | --- | --- | --- | --- | --- | --- | --- | --- | --- | --- |
|  | **Pre** | **Post** | **Pre** | **Post** | **Pre** | **Post** | **Pre** | **Post** | **Pre** | **Post** |
| **Anti-RBD**  (BAU/mL) | **247.8**  (5.2-1357) | **1624.0**  (10.7-7269) | **2.1**  (0.5-18.0) | **3.8**  (0.2-33.6) | **1590**  (894.1-3976) | **8990**  (5457-11360) | **247.6**  (14.8-725.5) | **1219**  (283.3-3587) | **335.2**  (59.7-3768) | **1929**  (316.9-10376) |
| **T cells**  (pg/mL) | **44.0**  (3.2-185.3) | **112.6**  (14.4-350.9) | **81.9**  (5.8-321.7) | **166.9**  (29.3-431.7) | **79.2**  (11.9-391.8) | **152.4**  (62-413.9) | **8.5**  (1.1-54.1) | **35.8**  (0.9-170.4) | **38.9**  (0.1-170.4) | 113.4  (4.2-323.0) |

**B: ID patients**

|  | **All**  **median (IQR)** | | **RTX**  **median (IQR)** | | **Other**  **median (IQR)** | |
| --- | --- | --- | --- | --- | --- | --- |
|  | **Pre** | **Post** | **Pre** | **Post** | **Pre** | **Post** |
| **Anti-RBD**  (BAU/mL) | **7.6**  (1.0-128.3) | **30.5**  (2.2-1458) | **2.2**  (0.1-7.6) | **3.7**  (0.5-30.5) | **279.3**  (56.6-949.3) | **1599**  (575.6-4052) |
| **T cells**  (pg/mL) | **74.8**  (28.1-233) | **175.5**  (60.1-595.2) | **37.4**  (19.7-259) | **289.2**  (76.8-531.5) | **76.8**  (44.8-171.7) | **153.9**  (25.1-690.8) |

**C: ST patients**

|  | **All**  **median (IQR)** | | **ChTX**  **median (IQR)** | | **ImmTX**  **median (IQR)** | | **TarTX**  **median (IQR)** | |
| --- | --- | --- | --- | --- | --- | --- | --- | --- |
|  | **Pre** | **Post** | **Pre** | **Post** | **Pre** | **Post** | **Pre** | **Post** |
| **Anti-RBD**  (BAU/mL) | **1546**  (810.4-4389) | **7632**  (3023-11360) | **1304**  (763.8-4206) | **7444**  (3613-11360) | **1508**  (811-9294) | **6937**  (4589-11360) | **1718**  (978.2-5568) | **7754**  (1985-11360) |
| **T cells**  (pg/mL) | **48.9**  (11.9-167.3) | **176**  (60.5-496) | **24.9**  (68.2-506) | **140**  (49.1-880.3) | **65.7**  (16.6-163.3) | **314**  (182.6-528) | **46.8**  (8.7-135.8) | **106**  (48.5-334.5) |

**The VAX4FRAIL Study Group**:

**Principal investigators** (alphabetical order): Giovanni Apolone (Fondazione IRCCS Istituto Nazionale dei Tumori di Milano); Alberto Mantovani (IRCCS Istituto Clinico Humanitas, Milano).

**Scientific Coordinators**: Massimo Costantini (Fondazione IRCCS Istituto Nazionale dei Tumori di Milano), Nicola Silvestris (Università degli Studi di Messina).

**Steering committee** (alphabetical order): Chiara Agrati (IRCCS Istituto per le Malattie Infettive Lazzaro Spallanzani, Roma); Giovanni Apolone (Fondazione IRCCS Istituto Nazionale dei Tumori di Milano); Fabio Ciceri (IRCCS Ospedale San Raffaele, Milano); Gennaro Ciliberto (IRCCS Istituto Nazionale Tumori Regina Elena, Roma); Massimo Costantini (Fondazione IRCCS Istituto Nazionale dei Tumori di Milano); Franco Locatelli (Università La Sapienza, Roma); Alberto Mantovani (IRCCS Istituto Clinico Humanitas, Milano); Fausto Baldanti (Fondazione IRCCS Policlinico San Matteo di Pavia); Aldo Morrone (Istituto Dermatologico San Gallicano IRCCS, Roma); Massimo Tommasino (IRCCS Istituto Tumori “Giovanni Paolo II”, Bari); Carlo Salvarani (Azienda USL-IRCCS Reggio Emilia); Nicola Silvestris (Università degli Studi di Messina); Giuseppe Lauri Pinter (Fondazione IRCCS Istituto Neurologico Carlo Besta, Milano); Antonio Uccelli (Ospedale Policlinico San Martino IRCCS, Genova); Pier Luigi Zinzani (IRCCS Azienda Ospedaliero-Universitaria di Bologna).

**Disease Groups**

1. Hematological malignancies Referent: Paolo Corradini (Fondazione IRCCS Istituto Nazionale dei Tumori, Milano);
2. Solid tumors Referent: Gennaro Ciliberto (IRCCS Istituto Nazionale Tumori Regina Elena, Roma);
3. Immunorheumatological diseases Referent: Carlo Salvarani (Azienda USL IRCCS Reggio Emilia);
4. Neurological diseases: Referent: Antonio Uccelli (Ospedale Policlinico San Martino IRCCS, Genova); Renato Mantegazza (Fondazione I.R.C.C.S Istituto Neurologico Carlo Besta (INCB), Milano).

**Immunological Group**

**Referents**: Chiara Agrati (IRCCS Istituto per le Malattie Infettive Lazzaro Spallanzani, Roma); Maria Rescigno (IRCCS Istituto Clinico Humanitas, Milano); Daniela Fenoglio (Ospedale Policlinico San Martino IRCCS, Genova);

**Participants**: Roberta Mortarini (Fondazione IRCCS Istituto Nazionale dei Tumori di Milano); Cristina Tresoldi (IRCCS Ospedale San Raffaele, Milano); Laura Conti, Chiara Mandoj (IRCCS Istituto Nazionale Tumori Regina Elena, Roma); Michela Lizier (IRCCS Humanitas Research Hospital, Rozzano, Milan); Stefania Croci (Azienda USL IRCCS Reggio Emilia); Fausto Baldanti (Fondazione IRCCS Policlinico San Matteo di Pavia); Vito Garrisi (IRCCS Istituto Tumori “Giovanni Paolo II”, Bari); Fulvio Baggi (Fondazione IRCCS Istituto Neurologico Carlo Besta, Milano); Tiziana Lazzarotto, Francesca Bonifazi (IRCCS Azienda Ospedaliero-Universitaria di Bologna); Fulvia Pimpinelli (Istituto Dermatologico San Gallicano IRCCS, Roma); Concetta Quintarelli (IRCCS Ospedale Pediatrico Bambino Gesù, Roma); Rita Carsetti (IRCCS Ospedale Pediatrico Bambino Gesù, Roma).

**INMI centralized laboratory** (INMI Lazzaro Spallanzani – IRCCS, Roma) (alphabetical order)

Enrico Girardi (Scientific Director), Aurora Bettini; Veronica Bordoni; Concetta Castilletti; Eleonora Cimini; Rita Casetti; Francesca Colavita; Flavia Cristofanelli; Massimo Francalancia; Simona Gili; Delia Goletti; Giulia Gramigna; Germana Grassi; Daniele Lapa; Sara Leone; Davide Mariotti; Giulia Matusali; Silvia Meschi; Stefania Notari; Enzo Puro; Marika Rubino; Alessandra Sacchi; Eleonora Tartaglia

**Clinical Task Force**

Paolo Corradini, Silvia Damian, Vincenzo Marasco, Filippo de Braud (Fondazione IRCCS Istituto Nazionale dei Tumori di Milano); Maria Teresa Lupo Stanghellini, Lorenzo Dagna, Francesca Ogliari, Massimo Filippi, Alessandro Bruno, Gloria Catalano, Rosamaria Nitti (IRCCS Ospedale San Raffaele, Milano); Andrea Mengarelli, Francesco Marchesi, Giancarlo Paoletti e Gabriele Minuti, Elena Papa (IRCCS Istituto Nazionale Tumori Regina Elena, Roma); Elena Azzolini, Luca Germagnoli, Carlo Selmi, Maria De Santis, Carmelo Carlo-Stella, Alexia Bertuzzi, Francesca Motta, Angela Ceribelli, Chiara Miggiano, Giulia Fornasa (IRCCS Humanitas Research Hospital, Rozzano, Milan); Fausto Baldanti, Sara Monti, Carlo Maurizio Montecucco (Fondazione IRCCS Policlinico San Matteo di Pavia); Aldo Morrone, Dario Graceffa (Istituto Dermatologico San Gallicano IRCCS, Roma); Maria Grazia Catanoso, Monica Guberti, Carmine Pinto, Francesco Merli, Franco Valzania (Azienda USL-IRCCS Reggio Emilia); Rosa Divella, Antonio Tufaro, Vito Garrisi, Sabina Delcuratolo, Mariana Miano (IRCCS Istituto Tumori “Giovanni Paolo II,” Bari; Carlo Antozzi, Silvia Bonanno Rita Frangiamore, Lorenzo Maggi (Fondazione IRCCS Istituto Neurologico Carlo Besta, Milano); Antonio Uccelli, Paolo Pronzato, Matilde Inglese, Carlo Genova, Caterina Lapucci, Alice Laroni, Ilaria Poirè (Ospedale Policlinico San Martino IRCCS, Genova); Marco Fusconi, Vittorio Stefoni, Maria Abbondanza Pantaleo (IRCCS Azienda Ospedaliero-Universitaria di Bologna).

**Statistical Committee**

Diana Giannarelli (IRCCS Istituto Nazionale Tumori Regina Elena, Roma).

**e-CRF and Monitoring Referent**

Valentina Sinno, Serena Di Cosimo (Fondazione IRCCS Istituto Nazionale dei Tumori di Milano).

**Project Managers of the Study**

**Referents:** Elena Turola, Azienda USL-IRCCS di Reggio Emilia.

**Participants**: Iolanda Pulice, Roberta Mennitto Fondazione IRCCS Istituto Nazionale dei Tumori, Milano); Stefania Trinca (IRCCS Ospedale San Raffaele, Milano); Giulia Piaggio (IRCCS Istituto Nazionale Tumori Regina Elena, Roma); Chiara Pozzi (IRCCS Humanitas Research Hospital, Rozzano, Milan); Irene Cassaniti (Fondazione IRCCS Policlinico San Matteo, Pavia); Alessandro Barberini (Istituto Dermatologico San Gallicano IRCCS, Roma); Arianna Belvedere (Azienda USL-IRCCS Reggio Emilia); Sabina Delcuratolo (IRCCS Istituto Tumori “Giovanni Paolo II,” Bari); Rinaldi Elena, Federica Bortone (Fondazione IRCCS Istituto Neurologico Carlo Besta, Milano); Maria Giovanna Dal Bello (Ospedale Policlinico San Martino IRCCS, Genova); Silvia Corazza (IRCCS Azienda Ospedaliero-Universitaria, Bologna).
